# Physiology-guided quantitative symptom analysis for gastroduodenal disorders

**DOI:** 10.1101/2023.06.07.23291112

**Authors:** Gabriel Schamberg, Chris Varghese, Emma Uren, Stefan Calder, Greg O’Grady, Armen A Gharibans, BSGM consortium

**Affiliations:** Department of Surgery, the University of Auckland, New Zealand; Alimetry Ltd, Auckland, New Zealand; Auckland Bioengineering Institute, The University of Auckland, New Zealand

## Abstract

**Background:** Current approaches to symptom-based classifications in gastroduodenal disorders are binary and substantially overlapping. We aimed to develop a standardized and quantitative system for classifying patient-level symptom profiles guided on physiological principles.

**Methods:** A large database (n = 787) of 4.5 h (30 min baseline; 4-h postprandial) Gastric Alimetry™ (Alimetry, NZ) recordings were used to identify, and quantify distinct symptom patterns based on established gastroduodenal physiology concepts. Tests comprised a standardized meal challenge and symptoms were simultaneously recorded at minimum 15 minute intervals using a 10-point likert scale with pictograms encoded in a validated digital App.

**Key Results:** Six symptom profiles were defined. The meal change metric was used to define ‘meal-induced’ and ‘meal-relieved’ symptom profiles, defined as an increase (+2) or decrease (−2) in the average symptom severity between the first post- and pre-prandial hours of recordings. The continuous profile was defined as a reduced range (<3; i.e., difference between the 95th and 5th percentile symptom severity), and thresholded to the 5th percentile of symptom severity being > 2. The symptom/amplitude correlation metric defined the ‘sensorimotor’ profile, thresholded when the correlation was >0.5. The symptom/amplitude time lag metric was used to define ‘activity-relieved’ and ‘post-gastric’ symptom profiles, defined as negative (< -0.25) or positive (>0.25) average difference between the cumulative distribution functions of the symptom and amplitude curves.

**Conclusions & Inferences:** Standardized quantification of symptom profiles in relation to a meal-stimulus and gastric amplitude offer a novel classification scheme based on gastroduodenal physiology.

## Introduction

Patient-reported symptoms serve as a foundation for clinical assessment and diagnosis of gastroduodenal disorders, primarily via the Rome-IV diagnostic questionnaires. However, significant limitations of current classifications include their binary consideration of disorder presence, significant overlap between classified gastroduodenal disorders, and inconsistent correlations with specific aetiologies or therapeutic outcomes.

Continuous symptom severity assessment in relation to a standardized meal stimulus offers an alternative approach that may be linked to pictograms and digital tools to enable specific, quantitative and comprehensive symptom profiling.(Tack et al. 2014; Sebaratnam et al. 2022) Average symptom severity has been shown to increase after meal ingestion in functional dyspepsia,(Stanghellini et al. 2016; Bisschops et al. 2008) and snapshot meal-related symptom severity profiles have been validated to correlate with longer-term daily symptoms and quality of life.(Kuwelker et al. 2021; Sebaratnam et al. 2022) However, a standardized system for classifying specific symptom profiles related to a meal change at the patient level has not yet been established. In addition, coupling postprandial symptom severity curves with specific measures of gastric physiology may provide synergistic clinical insights not available using either approach alone.(Gregory O’Grady et al. 2023; Wang et al. 2023)

The aim of this technical study was therefore to develop a standardized and quantitative system for classifying patient-level symptom profiles guided on physiological principles. The derived system was informed by results available from a large database of patients with chronic gastroduodenal symptoms evaluated using the Gastric Alimetry® system.

## Methods

Data collection was conducted in Auckland (New Zealand), Calgary (Canada), Leuven (Belgium) and Western Sydney (Australia). The study protocols were approved by The Auckland Health Research Ethics Committee (AHREC; AH1130), The University of Calgary Conjoint Health Research Ethics Board (REB19-1925),The UZ/KU Leuven Research Ethics Committee (S65541), and the Human Research Ethics Committee at Western Sydney (H13541). All data was collected in accordance with the guidelines and regulations of these committees. All subjects provided informed consent.

Gastric Alimetry is a new test of gastric function that enables simultaneous capture of high-resolution gastric myoelectrical activity and standardized symptom reporting on an 11-point likert scale (0 = none, 10 = most severe imaginable) using a validated app.(Sebaratnam et al. 2022) A complete description of the device and system is available elsewhere.(Gregory O’Grady et al. 2023; Gharibans et al. 2022) Using this system, we have compiled a database of 787 Gastric Alimetry tests from patients with chronic gastroduodenal symptoms, employing a standardized test format and meal, and with reference to established test reference intervals.(Gregory O’Grady et al. 2023; Varghese et al. 2023)

Employing this database, a standardized system was developed to quantify and classify a specific set of patient symptom profiles, and their relationships to simultaneously recorded gastric activity. This system was developed with the goal of facilitating quantitative analyses of the role of symptoms in clinical assessment of gastroduodenal disorders at scale. Robust metrics were introduced to jointly quantify physiological characteristics and symptom profiles into objective symptom phenotypes. Each characteristic, the associated metrics and phenotypes, and their clinical implications are discussed below with additional technical details in the **Supplementary Text**.

## Results

### Meal Response

The most studied characteristic of granular symptom data is the response of symptom severity to meal ingestion. Specifically, postprandial distress syndrome (PDS) is characterized by early satiation and/or excessive fullness, whereas epigastric pain syndrome (EPS), is associated with an increase or decrease in epigastric pain and/or burning following meal ingestion.(Stanghellini et al. 2016) Despite the importance of meal-related symptoms in gastroduodenal disorders, there is no agreed method for quantifying these changes. Therefore, to quantify the effect of meal ingestion on symptom severity, we define the *meal change* metric as the difference between the average symptom severities between the first hour postprandial and preprandial time windows. The *meal change* is thresholded to identify the **meal-induced** (*meal change* > 2) and **meal-relieved** (< -2) phenotypes.

### Symptom Persistence

It has previously been shown in patients with chronic nausea and vomiting disorders that those with normal Gastric Alimetry spectral analyses tend to have worse anxiety and/or depression than patients whose symptoms may be explained by gastric neuromuscular abnormalities.(Gharibans et al. 2022) It was further recently demonstrated that, of patients with normal BSGM spectral analyses, pre-meal high symptom severity and persistence of high symptoms throughout the test was a phenotype that was highly associated with anxiety and/or depression. (Wang et al. 2023) These results indicate that a high premeal symptom severity that persists through the test may be suggestive of disorders linked to the gut-brain axis. To quantify the symptom persistence, we define the *range* metric as the difference between the 95th and 5th percentile symptom severities. The **continuous** phenotype is identified by thresholding the *range* and the 5th percentile (*range* < 3 and 5th percentile > 2).

### Symptom-Amplitude Correlation

A recent review has identified that a subset of patients exhibit symptoms that are tightly time-synchronized with the gastric amplitude, indicating that these symptoms may have a sensorimotor component and may be suggestive of disorders linked to visceral hypersensitivity.(Greg O’Grady, Carbone, and Tack 2022; Gregory O’Grady et al. 2023) To rigorously quantify this relationship, we define the *symptom/amplitude correlation* as the correlation coefficient between the symptom severity curve and gastric amplitude curve. The *symptom/amplitude correlation* is thresholded to identify the **sensorimotor** (*correlation* > 0.5) phenotype.

### Symptom-Amplitude Time Lag

Patients may exhibit symptoms that occur either before the onset or after the conclusion of a physiological gastric meal response, suggesting that symptoms may be related to delayed onset of gastric mixing or a pathology distal to the stomach, respectively.(Gregory O’Grady et al. 2023) As a measure of the extent to which either of these patterns occur, we define the *symptom/amplitude time lag* as the average difference between the cumulative distribution functions of symptom and amplitude (−1 indicates all symptoms occurring before all gastric activity, and +1 all symptoms occurring after gastric activity). The *symptom/amplitude time lag* is thresholded to identify the **activity-relieved** (*lag* < -0.25) and **post-gastric** (> 0.25) phenotypes.

Based on the above scheme, the symptom metrics for the symptom severity curves profiled for nausea, bloating, upper gut pain, heartburn, and stomach burn were computed in 787 Gastric Alimetry tests performed on patients with chronic gastroduodenal symptoms. The proposed thresholds were used to phenotype all symptom curves, with the possibility for a symptom curve to have zero or multiple associated phenotypes. Symptom curves associated with each phenotype were visualized using the median curve and the associated interquartile range. For phenotypes relating symptom severity to gastric amplitude, the median (IQR) amplitude curves and average spectrograms for the patients with one or more symptom matching the phenotype are also shown. **Figures 1** and **2** illustrate that the proposed phenotypes produce distinct characterizations of the symptom severity curves and their relationships to time-synchronized gastric activity, and show their relative frequency of occurrence in a large cohort.

**Figure 1:**
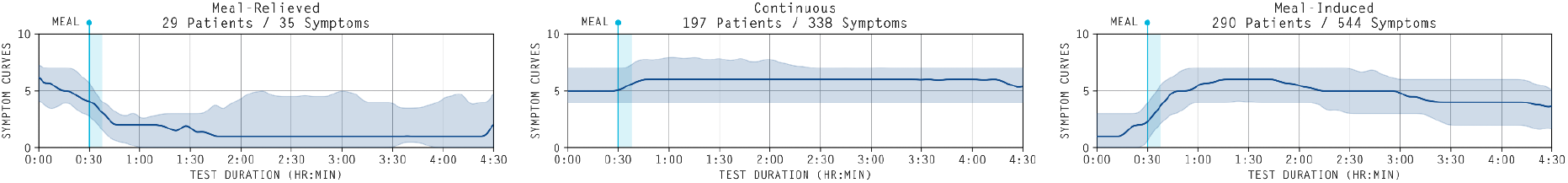
Median (IQR shaded) symptom severity curves for **meal-relieved, continuous**, and **meal-induced** symptoms. The number of patients with one or more symptoms matching the phenotype and the total number of symptoms matching the phenotype is indicated in the title.

**Figure 2:**
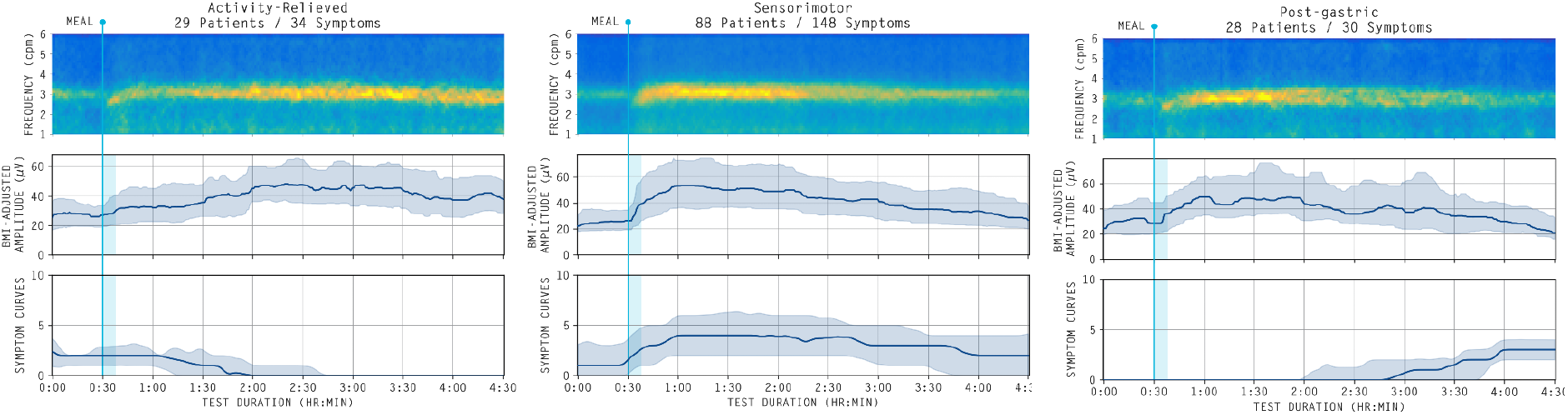
Average spectrograms and median (IQR shaded) BMI-Adjusted Amplitude curves and symptom severity curves for **activity-relieved, sensorimotor**, and **post-gastric** symptoms. The number of patients with one or more symptoms matching the phenotype and the total number of symptoms matching the phenotype is indicated in the title.

## Discussion

While continuously reported time-of-test symptoms have been explored in the literature, there remains no standardized method for characterizing various symptom profiles, and thus no structured framework for establishing their role in the assessment and diagnosis of gastroduodenal disorders.

Likewise, there has been no effort to study the relationship of symptom severity curves with concurrent myoelectrical activity of the stomach. Thus, we have introduced the first standardized approach to quantifying and classifying symptom profiles for relating continuous time-of-test symptoms to simultaneously recorded gastric activity. While the proposed approach was developed using the Gastric Alimetry system, it is applicable for any diagnostic test of gastroduodenal function applied in the context of a meal.

There are three key benefits to the proposed approach. First, we introduce the first formal metrics for quantifying the relationship between symptoms and simultaneously recorded gastric activity. We believe that such an approach will prove valuable for disentangling the complex interconnections that exist between patients’ experienced symptoms, gastroduodenal dysfunction, and disorders relating to the gut-brain axis.(Stanghellini et al. 2016; Black et al. 2020) Second, each of the proposed phenotypes is linked to a hypothesized physiological mechanism, enabling these phenotypes to guide further studies attempting to link symptom phenotypes with long-term outcomes to treatments and interventions. Lastly, by establishing a standardized and fully quantitative system for characterizing symptoms, the proposed approach can be applied at scale to assess the clinical relevance of each symptom profile in large patient populations.

With an established framework for performing quantitative symptom analysis and phenotyping, further clinical studies will need to be performed to establish the clinical significance of the proposed phenotypes and validate their hypothesized mechanisms. Specifically, observational studies can elucidate the relative prevalence of each phenotype in specific disease subgroups and establish support for the hypothesized mechanisms through comparisons with existing relevant patient data, such as questionnaires assessing stress, depression, quality of life, etc.(Gharibans et al. 2022) In the long term, studies evaluating the utility of symptom profiles for informing therapy and predicting patient outcomes will be necessary to establish their value in guiding diagnosis of gastroduodenal disorders.

In conclusion, we have introduced a framework for quantitatively assessing time-of-test symptoms as they relate to simultaneously recorded gastric myoelectrical activity. We anticipate that this framework will provide structure for further research into the role of symptoms in understanding gastroduodenal disorders, and ultimately could serve as a useful clinical tool for delivering targeted therapies to patients.

## Data Availability

Data will be made available upon reasonable request. Requests should be made to the corresponding author, Armen Gharibans.

## Acknowledgements

Funding was provided by the NZ Health Research Council Programme. Additional data collection was performed by clinical research assistants.

## Supplementary Text - Technical Details

### Preprocessing

During a Gastric Alimetry test, patients are able to update their symptom severities at any point, with reminders sent every 15 minutes. Before performing any quantitative analysis on a symptom severity curve, the curve is linearly interpolated to have one value per minute, such that it matches the timescale of the concurrently recorded gastric amplitude curve.

### Symptom Only Phenotypes

The **meal-induced, meal-relieved**, and **continuous** phenotypes are computed for a given symptom severity curve independently of the corresponding gastric amplitude.

The **meal-induced** and **meal-relieved** phenotypes are determined by thresholding the difference between the average symptom severity in the entire preprandial period (∼30 minutes) and the average symptom severity in the first hour following meal completion (difference > 2 and < -2, respectively).

The **continuous** phenotype is determined by applying a threshold to the range (the difference between 95th and 5th percentiles of a given curve) and the 5th percentile. Thresholding the range (< 3) ensures that the symptom severity is not changing significantly throughout the test, while thresholding the 5th percentile (> 2) ensures that the symptom is persisting at a non-negligible severity (i.e., persistently low symptoms should not be deemed **continuous**). The 95th and 5th percentiles are used in place of the maximum and minimum to ensure robustness to “misclicks” by the patient. For example, if a patient accidentally updates a symptom to be very low and immediately reverses it, that would affect the minimum but not the 5th percentile.

### Symptom/Amplitude Association Phenotypes

The **sensorimotor, activity-relieved**, and **post-gastric** phenotypes are determined by comparing the symptom curves to their associated time-synchronized gastric amplitude curves. Since each patient reports multiple symptoms, a single gastric amplitude curve will be compared to five different symptom severity curves. It is possible that a gastric amplitude curve will have missing values that have been automatically removed by the Gastric Alimetry Algorithm due to excessive movement artifacts. In such cases, the corresponding time points are also removed from the symptom severity curves, as the two curves cannot be compared where there is data missing from one. Additionally, all three of these phenotypes are only applied to cases where there is sufficient variation in both the amplitude and symptom severity curves. For example, if a patient reports changes by a maximum of one point on the 11-point likert scale, this may not be considered a significant enough change in symptoms to evaluate its relationship to gastric activity. Formally, we only identify symptom/amplitude association phenotypes when the standard deviation of the amplitude curve is > 10 μV and the standard deviation of the symptom severity curve is > 0.5.

The **sensorimotor** phenotype is determined by thresholding the correlation coefficient between the gastric amplitude and symptom severity curve (> 0.5). To account for the potential delay between the experience of symptoms and reporting of symptoms, this correlation coefficient is calculated as the maximum correlation coefficient allowing for a +/- 10 minute shift of the relative timing of the curves. Specifically, the correlation coefficient is calculated 21 times (shift = -10 min, …, 0 min, …, +10 min), with the maximum correlation coefficient compared to the threshold.

The **activity-relieved** and **post-gastric** phenotypes are determined by comparing the average difference between cumulative distribution functions (CDFs) associated with the amplitude and symptom severity curves (difference < -0.25 and > 0.25, respectively). The cumulative distribution function for each curve serves to represent where in the test the amplitude or symptom burden is concentrated. The steps to calculate the CDF for a given curve are: (1) normalize the curve so that it sums to one, (2) take the cumulative sum of the normalized curve. As a result, the CDF is a function that is increasing from zero at the beginning of the test to one at the end of the test, with the value at each point in time representing the proportion of the total amplitude or symptom burden that has already occurred. For example, if a particular symptom occurs primarily at the beginning of the test, the CDF will start at zero, and quickly rise to be close to one. As a result, the average (across time) difference between the CDFs for two curves will be positive if the concentration of gastric amplitude is earlier than that of the symptoms and negative if it is later.

## Notes

**Conflicts of Interest** A.G. and G.O. hold grants and intellectual property in the field of GI electrophysiology and are members of the University of Auckland spin-out companies: The Insides Company (G.O.), and Alimetry (G.S., E.U., S.C., G.O., and A.G.). The remaining authors have no relevant conflicts to declare.

### Competing Interest Statement

A.G. and G.O. hold grants and intellectual property in the field of GI electrophysiology and are members of the University of Auckland spin-out companies: The Insides Company (G.O.), and Alimetry (G.S., E.U., S.C., G.O., and A.G.). The remaining authors have no relevant conflicts to declare.

### Author Declarations

Ethics committee of the University of Auckland gave ethical approval for this work. Ethics committee of the University of Calgary gave ethical approval for this work. Ethics committee of KU Leuven gave ethical approval for this work. Ethics committee of Western Sydney University gave ethical approval for this work.

## References

Bisschops, R., G. Karamanolis, J. Arts, P. Caenepeel, K. Verbeke, J. Janssens, and J. Tack. 2008. “Relationship between Symptoms and Ingestion of a Meal in Functional Dyspepsia.” Gut 57 (11): 1495–1503.

Black, Christopher J., Douglas A. Drossman, Nicholas J. Talley, Johannah Ruddy, and Alexander C. Ford. 2020. “Functional Gastrointestinal Disorders: Advances in Understanding and Management.” The Lancet 396 (10263): 1664–74.

Gharibans, Armen A., Stefan Calder, Chris Varghese, Stephen Waite, Gabriel Schamberg, Charlotte Daker, Peng Du, et al. 2022. “Gastric Dysfunction in Patients with Chronic Nausea and Vomiting Syndromes Defined by a Noninvasive Gastric Mapping Device.” Science Translational Medicine 14 (663): eabq3544.

Kuwelker, Saatchi, David O. Prichard, Kent Bailey, and Adil E. Bharucha. 2021. “Relationship between Symptoms during a Gastric Emptying Study, Daily Symptoms and Quality of Life in Patients with Diabetes Mellitus.” Neurogastroenterology and Motility: The Official Journal of the European Gastrointestinal Motility Society 33 (12): e14154.

O’Grady, Greg, Florencia Carbone, and Jan Tack. 2022. “Gastric Sensorimotor Function and Its Clinical Measurement.” Neurogastroenterology and Motility: The Official Journal of the European Gastrointestinal Motility Society 34 (12): e14489.

O’Grady, Gregory, Chris Varghese, Gabriel Schamberg, Stefan Calder, Peng Du, William Xu, Jan Tack, et al. 2023. “Principles and Clinical Methods of Body Surface Gastric Mapping: Technical Review.” Neurogastroenterology and Motility: The Official Journal of the European Gastrointestinal Motility Society, March, e14556.

Sebaratnam, Gabrielle, Nikita Karulkar, Stefan Calder, Jonathan S. T. Woodhead, Celia Keane, Daniel A. Carson, Chris Varghese, et al. 2022. “Standardized System and App for Continuous Patient Symptom Logging in Gastroduodenal Disorders: Design, Implementation, and Validation.” Neurogastroenterology and Motility: The Official Journal of the European Gastrointestinal Motility Society 34 (8): e14331.

Stanghellini, Vincenzo, Francis K. L. Chan, William L. Hasler, Juan R. Malagelada, Hidekazu Suzuki, Jan Tack, and Nicholas J. Talley. 2016. “Gastroduodenal Disorders.” Gastroenterology 150 (6): 1380–92.

Tack, J., F. Carbone, L. Holvoet, H. Vanheel, T. Vanuytsel, and A. Vandenberghe. 2014. “The Use of Pictograms Improves Symptom Evaluation by Patients with Functional Dyspepsia.” Alimentary Pharmacology & Therapeutics 40 (5): 523–30.

Varghese, Chris, Gabriel Schamberg, Stefan Calder, Stephen Waite, Daniel Carson, Daphne Foong, William Jiaen Wang, et al. 2023. “Normative Values for Body Surface Gastric Mapping Evaluations of Gastric Motility Using Gastric Alimetry: Spectral Analysis.” The American Journal of Gastroenterology 118 (6): 1047–57.

Wang, William Jiaen, Daphne Foong, Stefan Calder, Gabriel Schamberg, Chris Varghese, Jan Tack, William Xu, et al. 2023. “Gastric Alimetry® Improves Patient Phenotyping in Gastroduodenal Disorders Compared to Gastric Emptying Scintigraphy Alone.” medRxiv. https://doi.org/10.1101/2023.05.18.23290134.

